# A Phase 1: Safety and Immunogenicity Trial of an Inactivated SARS-CoV-2 Vaccine-BBV152

**DOI:** 10.1101/2020.12.11.20210419

**Authors:** Raches Ella, Krishna Mohan Vadrevu, Harsh Jogdand, Sai Prasad, Siddharth Reddy, Vamshi Sarangi, Brunda Ganneru, Gajanan Sapkal, Pragya Yadav, Priya Abraham, Samiran Panda, Nivedita Gupta, Prabhakar Reddy, Savita Verma, Sanjay Kumar Rai, Chandramani Singh, Sagar Vivek Redkar, Chandra Sekhar Gillurkar, Jitendra Singh Kushwaha, Satyajit Mohapatra, Venkat Rao, Randeep Guleria, Krishna Ella, Balram Bhargava

## Abstract

**Background:** BBV152 is a whole-virion inactivated SARS-CoV-2 vaccine formulated with a TLR 7/8 agonist molecule adsorbed to alum (Algel-IMDG).

**Methods:** We conducted a double-blind randomized controlled phase 1 clinical trial to evaluate the safety and immunogenicity of BBV152. A total of 375 participants were randomized equally to receive three vaccine formulations (n=100 each) prepared with 3 μg with Algel-IMDG, 6 μg with Algel-IMDG, and 6 μg with Algel, and an Algel only control arm (n=75). Vaccines were administered on a two-dose intramuscular accelerated schedule on day 0 (baseline) and day 14. The primary outcomes were reactogenicity and safety. The secondary outcomes were immunogenicity based on the anti-IgG S1 response (detected with an enzyme-linked immunosorbent assay [ELISA] and wild-type virus neutralization [microneutralization and plaque reduction neutralization assays]). Cell-mediated responses were also evaluated.

**Results:** Reactogenicity was absent in the majority of participants, with mild events. The majority of adverse events were mild and were resolved. One serious adverse event was reported, which was found to be unrelated to vaccination. All three vaccine formulations resulted in robust immune responses comparable to a panel of convalescent serum. No significant differences were observed between the 3-μg and 6-μg Algel-IMDG groups. Neutralizing responses to homologous and heterologous SARS-CoV-2 strains were detected in all vaccinated individuals. Cell-mediated responses were biased to a Th-1 phenotype.

**Conclusions:** BBV152 induced binding and neutralising antibody responses and with the inclusion of the Algel-IMDG adjuvant, this is the first inactivated SARS-CoV-2 vaccine that has been reported to induce a Th1-biased response. Vaccine induced neutralizing antibody titers were reported with two divergent SARS-CoV-2 strains. BBV152 is stored between 2°C and 8°C, which is compatible with all national immunization program cold chain requirements. Both Algel-IMDG formulations were selected for the phase 2 immunogenicity trials. Further efficacy trials are underway.

Clinicaltrials.gov: NCT04471519

## Introduction

Severe acute respiratory syndrome coronavirus 2 (SARS-CoV-2), a novel human coronavirus(1), has spread globally. To date, 180 vaccine candidates are being developed to prevent coronavirus disease 2019 (COVID-19) (2). The virus strain (NIV-2020-770) isolated from a COVID-19 patient and sequenced at the Indian Council of Medical Research-National Institute of Virology (NIV) was provided to Bharat Biotech. Bio-Safety Level-3 manufacturing facilities and a well-established Vero cell manufacturing platform (with proven safety in other licensed live and inactivated vaccines) aided in the rapid development of BBV152 (3-7). BBV152 is a whole-virion inactivated SARS-CoV-2 vaccine adjuvanted with Algel and a TLR 7/8 agonist.

Preclinical studies in mice, rats, and rabbits and live viral challenge protective efficacy studies in hamsters and nonhuman primates aided in the clinical development of BBV152 (8-10). Here, we report the interim findings from the Phase 1 placebo-controlled randomized double-blind trial on the safety and immunogenicity of three different formulations of BBV152.

## Methods

### Trial Design and Participants

This is an adaptive randomized double-blind multicenter phase 1 trial to be seamlessly followed by a phase 2 trial to evaluate the safety, reactogenicity, tolerability, and immunogenicity of the whole-virion inactivated SARS-CoV-2 vaccine (BBV152) in healthy male and nonpregnant female volunteers. The participants were aged between 18 and 55 years at the time of enrollment. Participants were screened for eligibility based on their health status, including their medical history, laboratory findings, vital signs, and physical examination results, and were enrolled after providing signed and dated informed consent forms. Participants who tested positive for COVID-19 at screening by either the nucleic acid test or serology were excluded from the trial. Details of the inclusion and exclusion criteria can be found in the **Protocol**.

Participants were assigned a computer-generated randomization number. After their eligibility was determined, participants were randomized to four groups: 3 μg with Algel-IMDG, 6 μg with Algel-IMDG, 6 μg with Algel, and an Algel-only control arm. A two-dose intramuscular regimen was adopted with a 14-day interval. The trial was conducted across 11 sites in 9 states in India. The trial was approved by the National Regulatory Authority (India) and the respective Ethics Committees and was conducted in compliance with all International Council for Harmonization (ICH) Good Clinical Practice guidelines.

### Trial Vaccine

BBV152 (manufactured by Bharat Biotech) is a whole-virion, inactivated SARS-CoV-2 vaccine. The candidates were formulated with two adjuvants: Algel (alum) and Algel-IMDG, an imidazoquinoline class molecule (TLR7/TLR8 agonist abbreviated as IMDG) adsorbed onto Algel. Three vaccine formulations were prepared as follows: 3 μg with Algel-IMDG, 6 μg with Algel-IMDG and 6 μg with Algel. The placebo group contained only a sterile phosphate-buffered solution and Algel. The vaccine was provided as a sterile liquid that was injected through the intramuscular route at a volume of 0.5 mL/dose in a two-dose regimen with a 14-day interval. This accelerated schedule was chosen given the context of the ongoing pandemic. Both the vaccine and control were stored between 2°C and 8°C.

### Trial Procedures

Two doses of the BBV152 vaccine were administered at a volume of 0.5 mL/dose intramuscularly (deltoid muscle) on days 0 and 14. The follow-up visits were scheduled on days 7, 28, 42, 104, and 194. The study was performed in a dose-escalation manner wherein after completing vaccination in the first 50 participants with 3 μg with Algel-IMDG (the lowest antigen concentration) and the placebo, the participants were monitored for seven days for safety. Based on the independent Data Safety Monitoring Board (DSMB) recommendation, the trial was allowed to continue with enrollment of the remaining participants into all groups.

### Blinding

The appearance, color, and viscosity were identical across all treatment and control formulations. Participants, investigators, study coordinators, study-related personnel, and the sponsor were blinded to the treatment group allocation (excluding an unblinded CRO, who was tasked with the dispatch and labeling of vaccine vials and the generation of the master randomization code). Blinding was maintained using the randomization code.

### Safety Assessments

The primary safety outcome was the number and percentage of participants with solicited local and systemic reactogenicity within two hours, 7 days, 14 days, and 28 days after vaccination. No analgesics were given to participants before or after vaccination. Laboratory values (serum chemistry and hematology) were compared between the prevaccination (day 0) and postvaccination (day 28) visits.

Participants were observed for 2 hours postvaccination to assess the reactogenicity and were instructed to record the local and systemic reactions within seven days (days 0 to 7 and days 14 to 21) postvaccination using memory aids. Adverse events were graded according to the severity score (mild, moderate, or severe) and whether they were related or not related to the investigational vaccine, as detailed in the **Protocol**. Expected local reactogenicity included pain, tenderness, redness, erythema, swelling, induration, and systemic adverse events, including fever, fatigue/malaise, myalgia, body aches, headache, vomiting, anorexia, chills, rash, and diarrhea.

### Immunogenicity Assessments

Anti-IgG responses against the spike (S1) protein, receptor-binding domain (RBD), and nucleocapsid (N) protein of SARS-CoV-2 were assessed by enzyme-linked immunosorbent assay (ELISA) and are expressed as geometric mean titers (GMTs). Neutralizing antibody titers were evaluated by wild-type virus neutralization assays, namely, (i) a microneutralization assay (MNT_50_) and (ii) a plaque-reduction neutralization test (PRNT_50_), independently at Bharat Biotech and NIV. Details of these assays are provided in the **Supplementary Appendix**. To establish interlaboratory comparability, a subset of randomly selected serum samples (n=50) was analyzed by MNT_50_ at NIV and Bharat Biotech. Additionally, three challenge strains were utilized for PRNT_50_ at NIV: the BBV152 strain NIV-2020-770 – homologous, and two heterologous strains from the O Clade (NIV-Q111 and NIV-Q100). The NIV-2020-770 strain contains the D614G mutation.

To compare vaccine-induced responses to natural SARS-CoV-2 infections, 41 convalescent serum samples (collected within 1-2 months after nucleic acid test-based diagnosis) were tested by MNT_50_. These serum samples were collected from both symptomatic (n=25) and asymptomatic (n=16) COVID-19 patients at a regional hospital in Hyderabad. Seroconversion (SCR) rates were defined based on titers remaining ≥4-fold above baseline. All serum samples were analyzed in a blinded manner at Bharat Biotech and NIV.

Intracellular cytokine staining and ELISpot assays were used to assess T-cell responses. Peripheral blood mononuclear cells (PBMCs) were collected from a subset of participants (for interferon-gamma [IFN-γ]). These assays were performed at Bharat Biotech and Indoor Biotechnologies, India. The details of all assay methods can be found in the **Supplementary Appendix**.

### Statistical Analysis

The sample size was large to enable the immunogenicity comparisons among the groups, ensuring a high statistical power. The exact binomial calculation was used for the confidence interval estimation of proportions. The chi-square test or Fisher’s exact test was used to test differences in proportions. Confidence interval estimation for the geometric mean titer (GMT) was based on the log_10_ (titer) and the assumption that the log_10_ (titer) was normally distributed. A comparison of GMTs was performed with t-tests on the means of the log_10_ (titer). Significance was set at p < 0.025 (1-sided) or p < 0.05 (2-sided). No formal adjustment for multiple comparisons was planned. This preliminary report contains results regarding immunogenicity (captured on days 0 to 28) and safety outcomes (captured on days 0 to 42). Sample size estimation was performed using PASS 13 software (Number Cruncher Statistical Systems, USA), and descriptive and inferential statistics were performed using SAS 9.2.

## Results

Among the 897 potential participants screened between Jul 13 and Jul 30, 2020, 375 participants were randomized. Among the 522 initially screened individuals who were excluded, 70 and 63 participants were found to be positive for SARS-CoV-2 with the nucleic acid test and serology, respectively (**Figure 1**). Among the enrolled participants, 100 each were randomized into the three vaccine groups, and 75 were randomized into the control arm. The demographic characteristics of the participants are presented in Table S1 of **Supplementary Appendix**.

**Figure 1:**
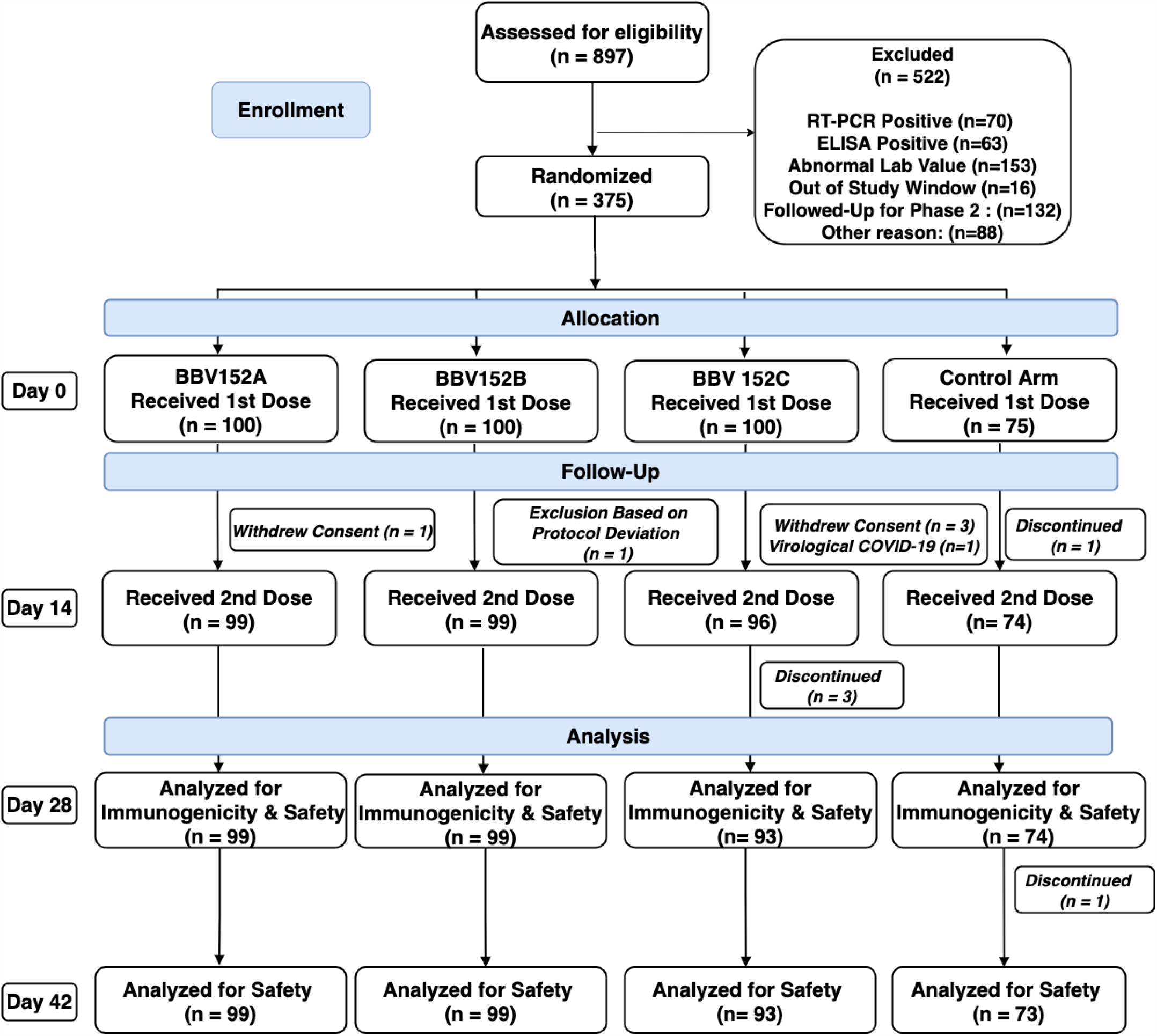
CONSORT Flow Diagram. Other exclusions (n=88) were unable to contact the participant for vaccination and withdrawal of consent. The study was performed in a dose-escalation manner wherein after completing vaccination in the first 50 participants with 3 μg with Algel-IMDG (the lowest antigen concentration) and the control (randomisation ratio 4:1); the participants were monitored for seven days for safety. Based on the independent Data Safety Monitoring Board (DSMB) reviewal of blinded safety data, the trial was allowed to continue with enrollment of the remaining participants into all groups.

### Safety

After the first vaccination, local and systemic adverse events were predominantly mild or moderate in severity and resolved rapidly, without any prescribed medication. A similar trend was observed after the second vaccination (**Figure 2**). Pain at the injection site was the most common local adverse event in the Algel-IMDG groups. The distribution of local and systemic AEs was equal among the vaccine treatment groups when compared to the control arm (**Figure 2**). Biochemical, hematological, and urine parameters outside of the normal ranges had no corroborating clinical manifestations (Table S11 in the **Supplementary Appendix**).

**Figure 2:**
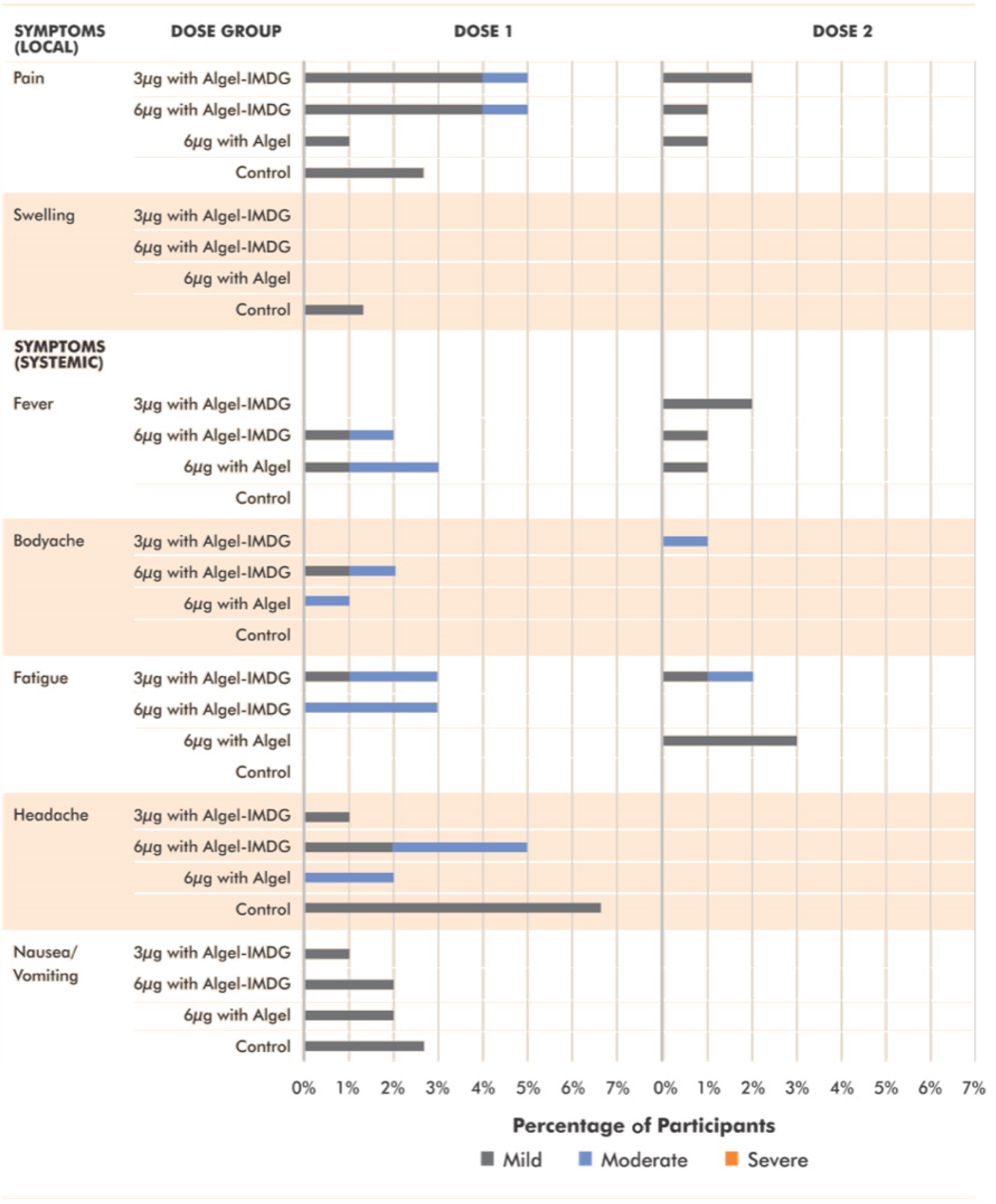
Solicited adverse events. The groups received the following: 3 μg with Algel-IMDG, 6 μg with Algel-IMDG, 6 μg with Algel, and Algel only as a control. Data are shown as the number of participants who experienced an event (%) after receiving either Dose 1 (0-7) or Dose 2 (14-21 days). The grading scale for most adverse events was based on the FDA guidance document for Toxicity Grading Scale for Healthy Adult and Adolescent Volunteers Enrolled in Preventive Vaccine Clinical Trials. For those adverse events where grading was not mentioned in the FDA guidance document, we have used the Common Terminology Criteria for Adverse Events (CTCAE) grading.

One serious adverse event was reported in the 6 μg with Algel group. The participant was screened on July 25^th^ and vaccinated on July 30^th^. Five days later, the participant reported symptoms of COVID-19 and was found to be positive for SARS-CoV-2 (by a nucleic acid test). The symptoms were mild in nature, but the patient was admitted to the hospital on August 15^th^. The participant was discharged on August 22^nd^ following a negative nucleic acid result. The event was not causally associated with the vaccine. No other symptomatic SARS-CoV-2 infections were reported between day 0 and 75.

### Immune Responses

#### Binding Antibody Titers

Anti-IgG titers (GMTs) to all epitopes (S1 protein, RBD, and N protein) increased rapidly after the administration of both doses (**Figure 3A-C)**. Both 3 μg and 6 μg with Algel-IMDG groups reported comparable anti-S1 protein, -RBD, and -N protein GMTs, adding to the dose-sparing effect of the adjuvant. Binding antibody titers to the whole virion inactivated antigen are highlighted in Figure S1 of the **Supplementary Appendix**. The isotyping ratios (IgG1/IgG4) were above 1 for all vaccinated groups, which was indicative of a Th1 bias (**Figure 3D)**.

**Figure 3:**
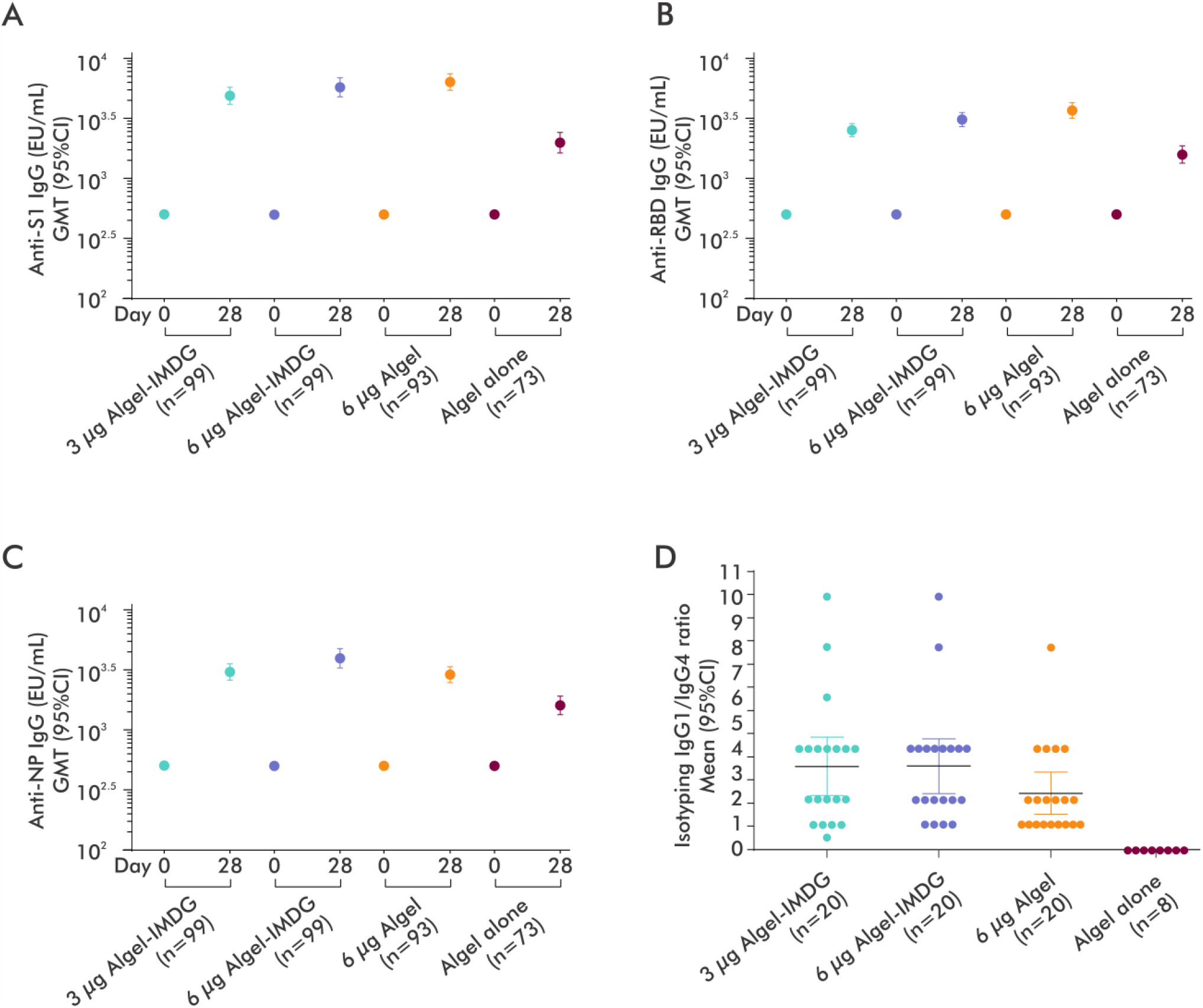
SARS-CoV-2 Antibody Responses (Anti S1, RBD, and N IgG) Shown are geometric mean reciprocal end-point enzyme-linked immunosorbent assay (ELISA) results at baseline (day 0) and 2 weeks after the second vaccination (day 28) for the 3 μg (n=99) and 6 μg (n=99) with Algel-IMDG groups, the 6 μg with Algel group (n=93), and the Algel-only control arm (n=73). Panels are segregated based on IgG titers against Anti-S1 (Panel A), Anti-RBD (Panel B), Anti-N (Panel C), and the Anti-S1 IgG1/IgG4 ratio (Panel D). In Panels A-C, dots and horizontal bars represent the geometric means and 95% CI, respectively. In Panel D, the isotyping ratio was calculated as IgG1/IgG4, and dots and horizontal bars represent the means and 95% CI, respectively.

#### Neutralizing Antibody Titers

The proportions of participants who experienced seroconversion (after the second dose) were 87.9%, 91.9%, 82.8% in the 3 μg with Algel-IMDG, 6 μg with Algel-IMDG, and 6 μg with Algel groups, respectively (**Figure 4A**). Seroconversion (at day 28) in the control arm was reported in 6 (8%) participants, suggestive of a high degree of ongoing infection. The post-second-dose GMTs (MNT_50_) in the three vaccine arms were 61.7, 66.4, and 48.0 in the 3 μg with Algel-IMDG, 6 μg with Algel-IMDG and 6 μg with Algel groups, respectively. Responses in the Algel-IMDG groups were noticeably higher than that in the 6 μg with Algel group, although the differences were not statistically significant. The vaccine-induced responses were comparable to those observed in the convalescent serum collected from patients who had recovered from COVID-19 (**Figure 4B**). The proportions of participants who experienced seroconversion analyzed by PRNT_50_ (after the second dose) were 93.4%, 86.4%, 86.6% in the 3 μg with Algel-IMDG, 6 μg with Algel-IMDG, and 6 μg with Algel groups, respectively. (**Figure 4C**). Randomly selected serum samples from day 28 were analyzed at NIV with homologous and heterologous strain challenges (**Figure 4D**).

**Figure 4:**
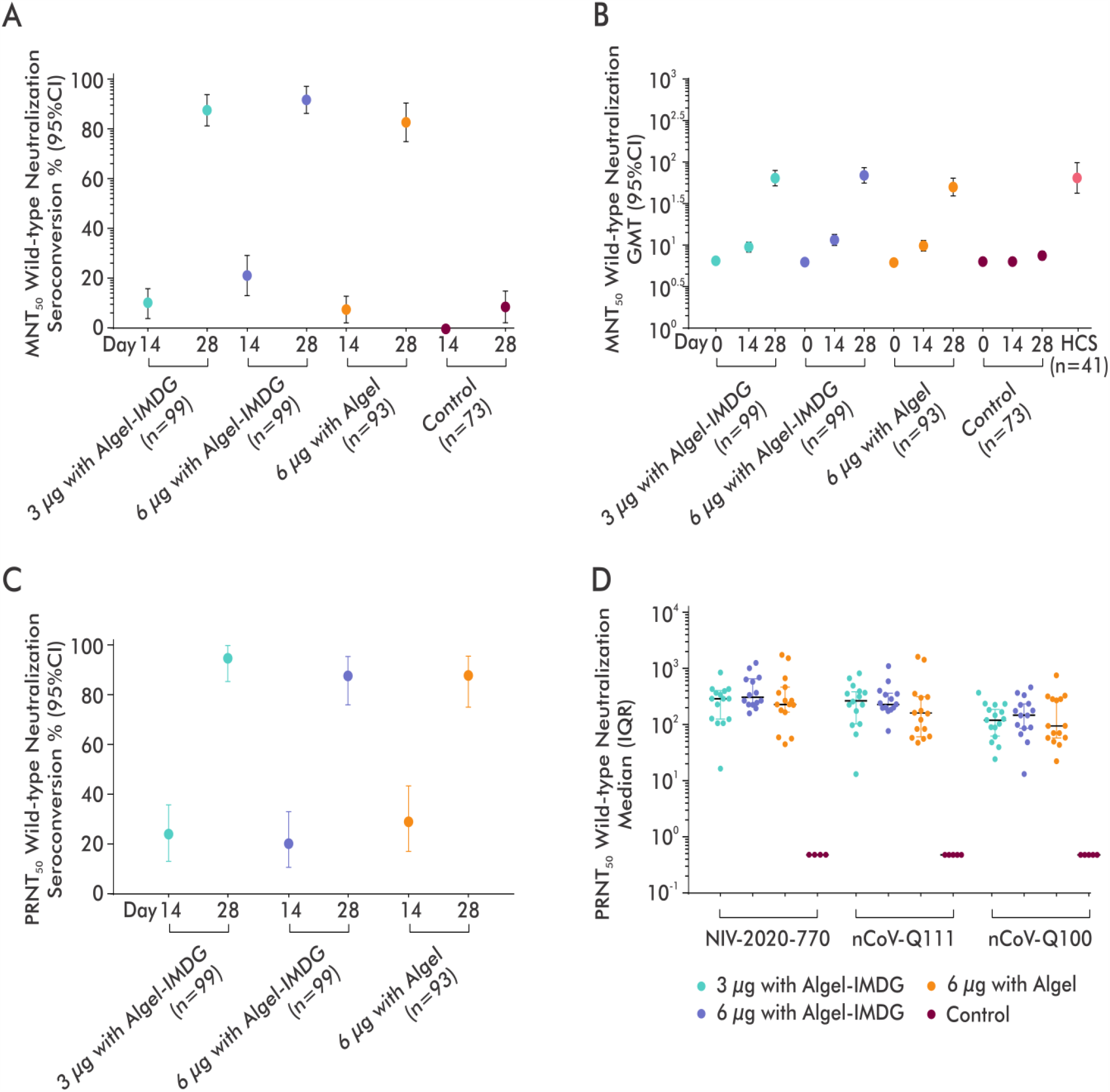
SARS-CoV-2 Neutralizing Antibody Responses. Shown are geometric mean titers of the wild-type SARS-CoV-2 microneutralization assay (MNT_50_) at baseline (day 0), 2 weeks after the first vaccination (day 14), and 2 weeks after the second vaccination (day 28) for the 3 μg and 6 μg with Algel-IMDG groups, the 6 μg with Algel group, and the Algel-only control arm. Seroconversion rates were defined by the proportion of titers remaining ≥4-fold above baseline. The dots and horizontal bars represent the SCR and 95% CI, respectively (Panel A). In Panel B, dots and horizontal bars represent the geometric means and 95% CI, respectively. The human convalescent serum (HCS) panel included specimens from PCR-confirmed symptomatic/asymptomatic COVID-19 participants obtained at least 30 days after diagnosis (41 samples for MNT_50_). In Panel C, seroconversion rates (analyzed wild-type SARS-CoV-2 plaque reduction neutralization assay [PRNT_50_] in the same immunogenicity cohort). The dots and horizontal bars represent the SCR and 95% CI, respectively). Randomly selected serum samples (n=15) from day 28 were analyzed by PRNT_50_ at the National Institute of Virology (NIV) for homologous (NIV-2020-770) and heterologous challenges (nCoV-Q11 and nCoV-Q100) (Panel D).

#### Cell-mediated Responses

IFN-γ ELISpot responses against SARS-CoV-2 peptides peaked at approximately 100-120 spot-forming cells per million PBMCs in all vaccinated groups on day 28. Both the Algel-IMDG groups elicited CD3^+^, CD4^+^, and CD8^+^ T-cells that were reflected in the IFN-γ production. There was minimal detection in the 6 μg with Algel group and the control arm (**Figure 5** and Figure S4 in the **Supplementary Appendix**).

**Figure 5:**
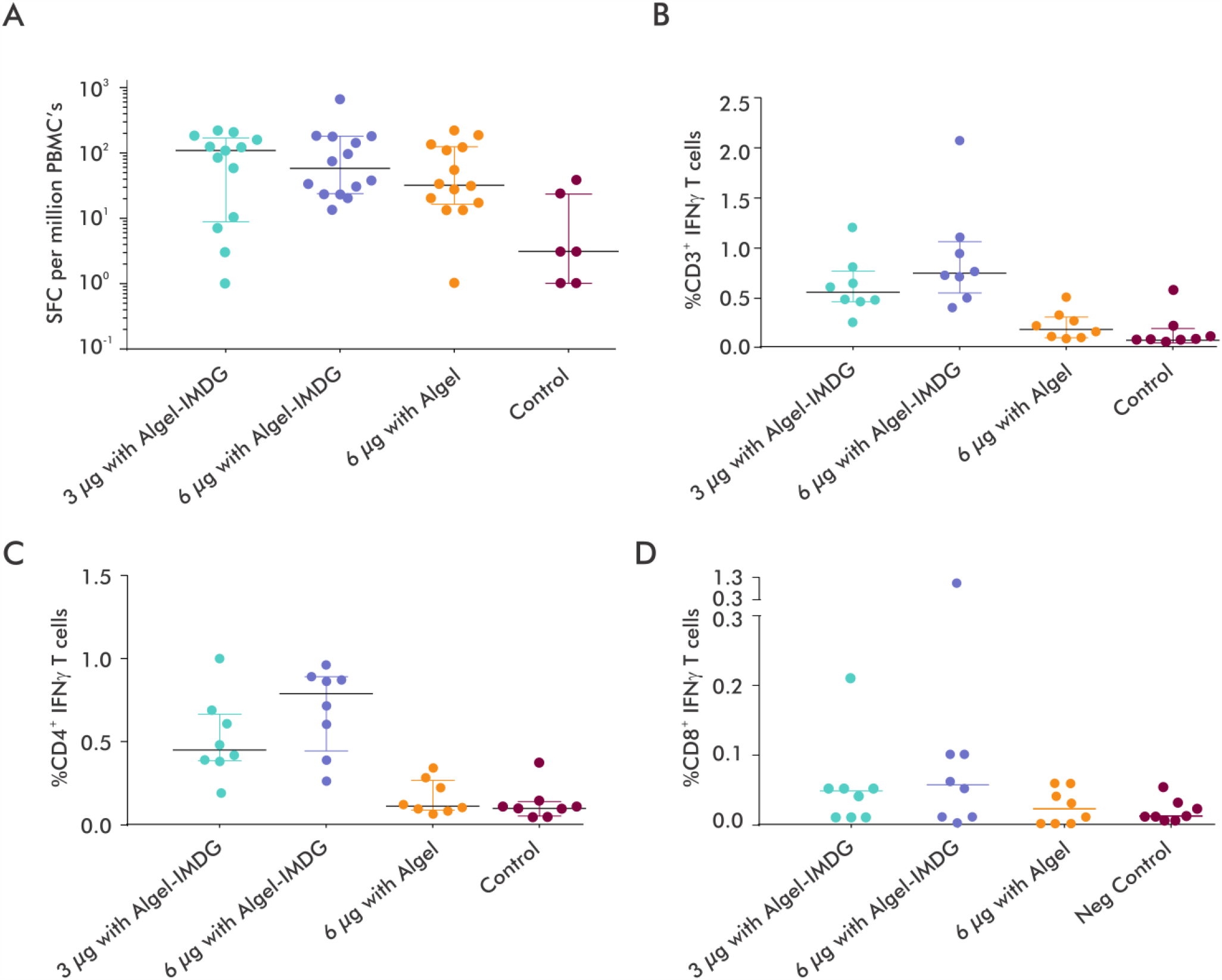
SARS-CoV-2 Cell-mediated Responses. Shown are the frequencies of antigen-specific *cell-mediated responses*, 2 weeks after the second vaccination (day 28) for the 3 μg and 6 μg with Algel-IMDG groups, 6 μg with Algel group, and the Algel-only control arm. Error bars show the median (IQR). *Interferon-gamma (IFN-γ) ELISpot response* (spot-forming cells [SFCs] per million peripheral blood mononuclear cells [PBMCs]) *to peptides spanning the SARS-CoV-2 spike and N proteins* (Panel A). Intracellular cytokine staining was used to assess the frequencies of antigen-specific CD3^+^, CD4^+^, and CD8^+^ T-cells (producing *IFN-γ*), as depicted in Panels B, C, and D, respectively.

## Discussion

We report the interim findings from this phase 1 clinical trial of BBV152, a whole-virion inactivated SARS-CoV-2 vaccine. The vaccine was well tolerated in all dose groups with no vaccine-related serious adverse events. Robust humoral and cell-mediated responses were observed in the Algel-IMDG recipients.

The most common adverse event was pain at the injection site, which resolved spontaneously. The overall incidence rate of local and systemic adverse events in this study was 10%-20% in all vaccine treated arms, which is noticeably lower than the rates for other SARS-CoV-2 vaccine platform candidates (11-15) and comparable to the rates for other inactivated SARS-CoV-2 vaccine candidates (11, 16).

One serious adverse event was reported in the 6 μg with Algel group. The participant was screened on July 25^th^ and vaccinated on July 30^th^. Five days later, the participant reported fever and headache (initially reported as a solicited adverse event), and on August 8^th^ was found to be positive for SARS-CoV-2 (by a nucleic acid test). The symptoms were initially mild in nature, which the onset of relapsing fever requiring admission to the hospital on August 15^th^. The participant had stable vitals (except body temperature) during hospitalization and did not require supplemental oxygen. The participant was discharged on August 22^nd^ following a negative nucleic acid result. The event was not causally associated with the vaccine. No other symptomatic SARS-CoV-2 infections were reported between day 0 and 75. However, follow-up of routine SARS-CoV-2 nucleic acid testing was not conducted on any scheduled or illness visit. We recognize that this study was conducted during a time of high ongoing degree of COVID-19 circulation, as evidenced by 8% of the participants in the placebo arm seroconverting with antibodies to SARS-CoV-2 (at day 28).

Whole-virion inactivated vaccines have been used for decades and have a well-established safety profile. Bharat Biotech manufactures several Vero cell-derived whole-virion inactivated licensed vaccines (5-7) and two investigational candidates (Zika and Chikungunya). We have accumulated safety data from 4,700 participants across phase 1-3 clinical trials and conducted pharmacovigilance reports on all licensed vaccines (approximately 100 million doses administered).

Whole-virion inactivated vaccines are mostly developed with Algel (alum) as the adjuvant. The response generated by alum is primarily Th2-biased, with the induction of strong humoral responses by neutralizing antibodies (17). A few animal studies of animal SARS-CoV-1 and MERS-CoV inactivated or vectored vaccines adjuvanted with alum have shown Th2 responses resulting in eosinophilic infiltration in the lungs (18-21). Complicating adverse events may be associated with the induction of weakly neutralizing or nonneutralizing antibodies that lead to antibody-dependent enhancement (ADE) or enhanced respiratory disease (ERD), thus prompting the attempt to develop SARS-CoV-2 that induce a CD4^+^ Th1 response with a minimal Th2 response (22-24).

Previous studies have shown that the Toll-like receptors (TLRs) play an integral role in bridging the innate and adaptive immune responses, leading to the differentiation of CD4^+^ T-cells into Th1 cells, which produce IFN-γ (25). Geeraedts et al. reported that the stimulation of TLR7 by an influenza whole-virion inactivated vaccine was a significant determinant of a greater immune response and Th1 polarization (26). Thus, it is imperative to develop such whole-virion inactivated vaccines with adjuvants that can synergistically contribute to the full potential. Algel-IMDG contains an imidaquizoquinoline class TLR7/8 agonist adsorbed to Algel. Preclinical studies on BBV152 adjuvanted with this molecule reported a Th1-biased response in mice (8). Furthermore, in a nonhuman primate and hamster live viral challenge study, Algel-IMDG formulations led to higher neutralizing antibodies, which may have resulted in improved upper and lower airway viral clearance (postchallenge) (9, 10).

BBV152 induced robust binding and neutralizing antibody responses that were similar to those induced by other SARS-CoV-2 inactivated vaccine candidates (11, 16). Here, we demonstrated that all vaccine formulations were Th1 skewed with IgG1/IgG4 ratios above 1. Furthermore, the Algel-IMDG formulations were associated with a significant increase in the frequency of CD4^+^ INF-γ^+^ T-cells when compared to the 6 μg with Algel formulation, which is indicative of a Th1 bias. Additionally, cell-mediated responses from other SARS-CoV-2 inactivated vaccine candidates have not been reported thus far.

This study has a few limitations. First, as this is an interim report, we are not reporting any data on the persistence of vaccine-induced antibody responses or safety outcomes. Second, the results reported here do not permit efficacy assessments. Third, the evaluation of safety outcomes requires more extensive Phase 3 clinical trials. Last, we evaluated an accelerated schedule (vaccination occurred 2 weeks apart) and did not include a routine schedule (vaccination occurring 4 weeks apart). The latter schedule is being evaluated in Phase 2 (27).

However, this study has several strengths. To ensure generalizability, this study was conducted with participants from diverse geographic locations and socioeconomic conditions, enrolling 375 participants across 11 hospitals. Despite the fact that enrollment occurred during a national lockdown, which led to several operational challenges, the overall participant retention rate was 97%. The sample size was intentionally large to enable the inference of meaningful conclusions regarding immunogenicity and safety.

BBV152 induced binding and neutralising antibody responses and with the inclusion of the Algel-IMDG adjuvant, this is the first inactivated SARS-CoV-2 vaccine that has been reported to induce a Th1-biased response. BBV152 is stored between 2°C and 8°C, which is compatible with all national immunization programs. Both Algel-IMDG formulations were selected for the phase 2 immunogenicity trials. Further efficacy trials are underway.

## Data Availability

This is an interim report of an ongoing clinical trial. This report was prespecified in the protocol.

## Acknowledgments

Our sincere thanks to the principal and co-principal investigators, study coordinators and health care workers that were involved in this study. We express our gratitude to Dr. Sivasankar Baalasubramaniam from Indoor Biotechnologies, Bangalore, who assisted with cell-mediated response analyses and Dr. Dipankar Das from Bharat Biotech, for binding antibody estimation. A special thanks to Drs. Arjun Dang and Leena Chatterjee of Dr. Dangs Lab, which was the central laboratory for clinical laboratory testing. Drs. Shashi Kanth Muni, Sapan Behera, Vinay Aileni, and Ms. Akhila Naidu of Bharat Biotech, participated in protocol design and clinical trial monitoring. This vaccine candidate could not have been developed without the efforts of Bharat Biotech’s manufacturing and quality control teams. All authors would like to express their gratitude for all frontline health care workers during this pandemic.

## Author Contributions

All listed authors meet the criteria for authorship set forth by the International Committee for Medical Editors and have no conflicts to disclose. J.H., B.G., P.Y., and G.S. led the immunogenicity experiments. K.M.V., P.S., and E.R. contributed to the analysis and manuscript preparation. S.R. was study coordinator and helped immensely with the protocol design and interim report generation. P.A., S.P., A.P., N.G., B.B., of NIV and ICMR, India, contributed various neutralizing antibody assays and participated in the writing of this manuscript. All principal investigators were involved with the scientific review of this manuscript.

